# Mitigating the transmission of infection and death due to SARS-CoV-2 through non-pharmaceutical interventions and repurposing drugs

**DOI:** 10.1101/2020.09.28.20202804

**Authors:** Chittaranjan Mondal, Debadatta Adak, Abhijit Majumder, Nandadulal Bairagi

## Abstract

The Covid-19 pandemic has put the world under immeasurable stress. There is no specific drug or vaccine that can cure the infection or protect people from the infection of coronavirus. It is therefore prudent to use the existing resources and control strategies in an optimal way to contain the virus spread and provide the best possible treatments to the infected individuals. Use of the repurposing drugs along with the non-pharmaceutical intervention strategies may be the right way for fighting against the ongoing pandemic. It is the objective of this work to demonstrate through mathematical modelling and analysis how and to what extent such control strategies can improve the overall Covid-19 epidemic burden. The criteria for disease elimination & persistence were established through the basic reproduction number. A case study with the Indian Covid-19 epidemic data is presented to visualize and illustrate the personal hygiene & safe distancing, and repurposing drugs. It is shown that India can significantly improve the overall Covid-19 epidemic burden through the combined use of NPIs and repurposing drugs though containment of spreading is difficult without serious community participation.

## 1. Introduction

The world has been passing through an extraordinary crisis period since late December 2019 due to an extraordinary pathogen Covid-19 [1]. This virus is extraordinary because our immune system is unable to fight against this virus, and it has stunning transmission capability from human-to-human. The crisis is extraordinary because it has not only caused a global medical crisis but also put the world into unprecedented economic and social crises [2]. The health care system throughout the globe is under tremendous stress, and the global economy is likely to face the deepest recession after the Great Depression of the 1930s [3].

The story began on the last day of 2019 when China informed the World Health Organization (WHO) about the outbreak of 2019-nCoV in the Wuhan City of its Hubei Province. The virus crossed the border of China and disseminated in 23 other countries including the USA, France, Spain, Italy, UK and India through international travellers in the first month of 2020 [4]. Understanding the seriousness of the infection, WHO declared 2019-nCoV outbreak as a public health emergency of international concern (PHIC) on January 30 for the sake of accelerated preparedness and better management to prevent further spread of infection to other countries. Unfortunately, this alert call was in vain and the virus spread to 200 countries as of March 11 when the WHO declared Covid-19 a global pandemic. As of August 27, 216 countries/territories are affected by this dreaded disease with more than 24.06 million positive cases and 0.82 million deaths globally [5].

The coronavirus spreads when an individual inhales the droplets containing virus [4]. Such contaminated droplets are produced during sneezing and coughing by an infected individual. Study shows that these droplets usually cannot travel more than 2 meters from its source and it is therefore advised to maintain a distance of six feet between two individuals to avoid an infection [6]. The number of suspended droplets, however, can be reduced through practice of good cough and sneeze etiquette, which reduces the transmission probability of infection from an infective to a susceptible. Using of face musk is one of the most effective nonpharmaceutical measures that protect individuals from inhaling of virus-carrying droplets and restricts transmission [7, 8]. An individual may also be infected if one touches a contaminated surface and then touches his/her mouth, nose and eyes [9]. This mode of transmission may be prevented by disinfecting the surfaces. Individual hygiene practice like frequent washing of hands by soap or sanitizer may effectively reduce the probability of getting the infection through surface contamination. Using of spectacles and sunglass may reduce the chances of contamination through eye [10, 11]. However, the most effective way to reduce large-scale contamination and community transmission is the lockdown [12]. To prevent human-to-human transmission of the coronavirus through contact, many countries have implemented nationwide or region-wide lockdown by closing academic institutions, offices, restaurants, community halls, social gathering and all modes of transportation. This mechanism has been proved to be effective in containing the transmission but unable to eradicate the disease in the absence of a vaccine [13]. For example, countries like China, South Korea and most of the European countries have been able to contain the infection but no country is able to eradicate this disease from its geographical region [14, 15, 16].

Till today, neither there is any specific drug that can cure the infection nor there is any vaccine that can protect people from SARS-CoV-2 [17, 18]. Covid-19 vaccine tracker [19] shows that at least 41 potential vaccines are currently in phase 1-3 trials. Scientists are also searching for specific drugs that can be used to cure SARS-CoV-2 infection from an individual body. Vaccine and drug development process is time-consuming and may take a long period for its availability in the market even after using a fast-track. During such pandemic situations, one can not wait for a vaccine or a new drug, rather it would be prudent to fight against the virus with the available weapons in hand. Until specific drugs and vaccines are available, some repurposing drugs may be effectively used to save the lives of many severely infected covid patients [20]. There are many benefits of using repurposing drugs [21, 22]. First, infected patients may be given some quick treatments with such drugs. Second, since these drugs have already gone through rigorous safety tests, it can be applied to the patients without going through the time-consuming safety trials. Third, the overall treatment cost is less as there is no expense for drug development. Existing drugs like dexamethasone, favipiravir and remdesivir are some of such drugs which have been proven to be useful in treating covid-19 patients. Dexamethasone is a corticosteroid used for multiple problems like arthritis, asthma, intestinal disorder etc. The WHO and the NIH (National Institute of Health) have recently approved its use for the treatment of acute Covid-19 patients. It has been shown through randomized clinical trials that the immunosuppressant dexamethasone can save the lives of critically ill (on ventilation) Covid-19 patients and can reduce the mortality rate by one-third [23, 24]. An RNA polymerase inhibitor remdesivir, a drug used for Ebola virus infection, has shown the prophylactic and therapeutic efficacy for patients with severe Covid-19 [25, 26]. In vitro and in vivo experiments confirmed the efficacy of the nucleotide prodrug remdesivir against coronaviruses [27]. Both the European Union [28] and the USA [29] governments have approved its use in the treatment of COVID-19. Another important treatment study that can give life to many seriously ill Covid-19 patients is convalescent plasma or immunoglobulins therapy [30]. This passive immunization therapy has the potentiality to improve the survival rate of covid patients [31]. Use of such repurposing drugs along with the non-pharmaceutical intervention strategies may be the best possible way for fighting against the ongoing pandemic in the absence of any vaccine and specific drug for this novel coronavirus. It is the objective of this work to demonstrate through mathematical modelling and analysis how and to what extent the NPIs and repurposing drugs can improve the overall Covid-19 epidemic burden.

Numerous mathematical models on Covid-19 pandemic have been appeared to forecast the future of the epidemic. These models reasonably address the epidemic burden of an affected country to guide the policymakers and the healthcare providers on preparedness. Fitting the data of China, Italy and France with a SIRD model, Fanelli and Piazza [32] showed that the kinetic parameter that represents the recovery rate remains same for many countries but the death rate of the respective countries are different. A higher dimensional transmission model was proposed in [33] to study the Covid-19 epidemic of Wuhan. In Chatterjee et al. [34], a simple SEIR compartmental model was simulated with Monte Carlo (MC) simulation technique to measure the effect of NPIs. An epidemic model was used to investigate the dynamics of SARS-CoV-2 with multiple transmission pathways by Yang and Wang [17]. They infer from the analytical and numerical results that the infection will remain endemic for a long time. A regression model for Covid-19 was used to estimate the final size and its peak time for China, South Korea, and the rest of the World [35]. In [36], a simple SIR model was used to estimate various epidemic parameters by fitting the epidemic data of the Republic of Korea. Nonlinear incidence was used in an epidemic model to show that lockdown can make significant delay to attain the epidemic peak but unable to eradicate the disease [37]. A simple iteration model, which uses only daily values of confirmed cases, was considered to forecast the covid positive cases for the United States, Slovenia, Iran, and Germany [38]. Pal et al. [39] explained the Covid-19 transmission dynamics during the unlock phase and the significance of testing with the help of a mathematical model. Simulating an SEIR model, Fang et al. [40] studied the impact of different control measures in the spreading of coronavirus. A computational model was used in [41] to assess the impact of face musk in the disease spreading of Covid-19. It is shown that universal use of face masks along with other non-pharmaceutical practices can significantly inhibit the disease progression.

In this paper, we have proposed a minimal epidemic model to capture the dynamics of observed data of detecting, recovered and death cases of any Covid-19 affected country. The model is then extended to include three control strategies to mitigate the transmission of Covid-19 and to reduce its related deaths. Human-to-human transmission of infection depends mainly on two things: the number of per capita daily contacts between susceptible and infectives, and the probability of disease transmission per effective contact [42]. As mentioned before, per capita daily contacts may be significantly reduced through lockdown, while the probability of virus transmission can be reduced by using face masks and other good hygiene practices. We, therefore, used these nonpharmaceutical interventions as two control measures. The third control is used to reduce the death rate of severely infected Covid-19 patients using repurposing drugs. The proposed epidemic model is then analyzed to determine the criteria for persistence and extinction of the disease. It is shown that repurposing drugs is very useful in saving lives and increasing the recovery rate. Through a case study, it is shown that India can significantly improve the overall Covid-19 epidemic burden through the combined use of NPIs and repurposing drugs and it is true for any other country. India can save lives of 4794 corona infected patients in the next one month (August 28 to September 26) if the repurposing drug has efficacy 0.3.

The rest of the paper is organized as follows. In Section 2, we present the mathematical model to be analyzed. Different mathematical results that determine the dynamics of the system are given in Section 3. A case study with Indian Covid-19 epidemic data is presented in Section 4. Various results are demonstrated there in relation to the proposed model. The paper ends with a discussion in Section 5.

## 2. Model construction

Here we consider five compartments, viz. susceptible (*S*), latent (*E*), home isolated (*I*_*h*_), hospitalized (*I*_*c*_) and recovered (*R*) classes, to classify the human population of a coronavirus affected country based on their health status. Thus, *N*(*t*) = *S* (*t*)+ *E*(*t*)+ *I*_*h*_(*t*)+ *I*_*c*_(*t*)+*R*(*t*) represents the total population of the country at any time *t*. As SARS-CoV-2 is a novel virus, every individual is assumed to be susceptible to this virus. After getting an infection, an individual joins the latent class, *E*. Latent individuals are detected through real-time polymerase chain reaction (RT-PCR) test and advised for home isolation. Some infected individuals under home isolation state (*I*_*h*_) may develop complicacy and are shifted to hospital for necessary treatment. Such critically infected individuals are denoted by *I*_*c*_. Critically ill Covid-19 patients join the class *R* after recovery or succumb to infection. It is further assumed that individuals join *S* class through birth at a rate *G*. The average per capita daily contacts of an infected individual is assumed to be *n* and the probability of disease transmission through contact between an infected individual and a susceptible individual is *p*. A common practice while writing the incidence term is to express the product of *n* and *p* as a single term, called the disease transmission coefficient or the force of infection [42]. We, however, express it explicitly because different NPIs affect differently on these parameters. Individuals of *E* class are identified as covid positive at a rate of δ and immediately join *I*_*h*_ class. A proportion ω of *I*_*h*_ class develops complicacy and are transferred to *I*_*c*_ class for the treatment. The parameters *v*_1_, *v*_2_ and *m*_1_, *m*_2_ are the recovery and death rates of *I*_*h*_ and *I*_*c*_ classes, respectively. There are reports that covid patients die in transit due to lack of hospital beds, unavailability of ambulance [43, 44, 45]. We, therefore, assume that some individuals of *I*_*h*_ class may also succumb to infection before they are shifted to *I*_*c*_ class. It is, however, true that most of the members in mildly infected class (*I*_*h*_) recover from the infection. Therefore, it would be justified to assume that ν_1_ >> ν_2_ and *m*_1_ << *m*_2_. The natural death rate of susceptible individuals is assumed to be σ, giving 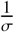 as the average life expectancy of the common people, and considered in all compartments. We also consider a death class *V* to represent the virulence of the disease (Covid-19 related death). Note that the individuals of the death class are not included to represent the total population. The dimension of the proposed model was kept low, though this simplified representation reflects the usual coronavirus infection management protocol followed in many countries including India. With these assumptions, we propose the following model for Covid-19 epidemic:

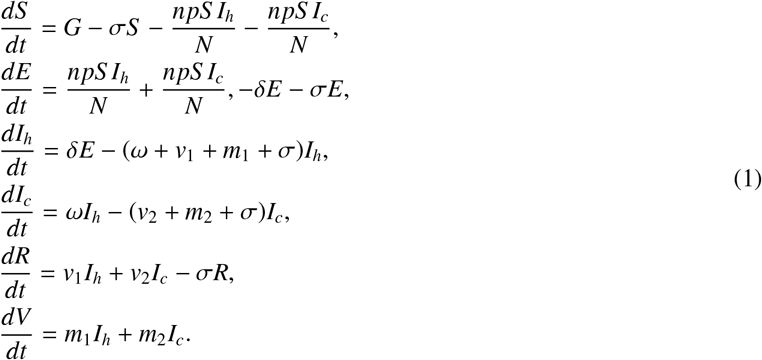

Those who are critically ill are assumed, for simplicity, to be unable to spread the infection further. This assumption may be too strict but simplifies the model to

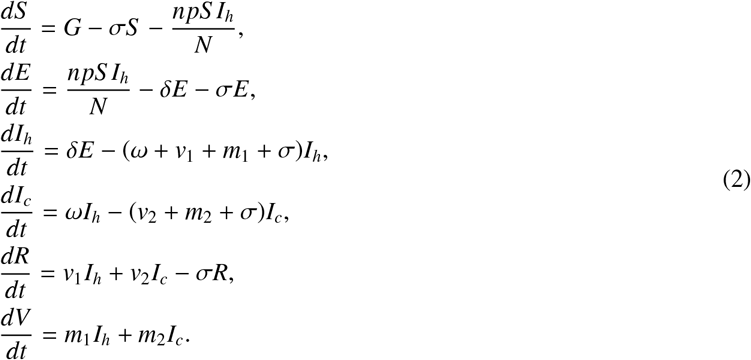

A schematic representation of the system (2) is given in Fig. 1. Parameters and variables are defined in Table 1.

**Table 1:** Variables and parameter with their definitions

| Variable | Description | Default value |
| --- | --- | --- |
| $S(t)$ | Susceptible human population at time $t$ | - |
| $E(t)$ | Latent individuals at time $t$ | - |
| $I_h(t)$ | Home isolated infected individuals at time $t$ | - |
| $I_c(t)$ | Hospitalized infected individuals at time $t$ | - |
| $R(t)$ | Recovered individuals at time $t$ | - |
| $V(t)$ | Virulence class individuals at time $t$ | - |
| Parameter | Description | Default value |
| $G$ | Constant input of susceptible individuals through birth | To be estimated |
| $\sigma$ | Natural death rate | To be estimated |
| $n$ | Average per capita daily contact | To be estimated |
| $p$ | Probability of disease transmission per contact | To be estimated |
| $\delta$ | Leaving rate from E class | To be estimated |
| $\omega$ | Transfer rate from $I_h$ class to $I_c$ class | To be estimated |
| $\nu_1$ | Recovery rate of $I_h$ class | To be estimated |
| $m_1$ | Death rate of $I_h$ class | To be estimated |
| $\nu_2$ | Recovery rate of $I_c$ class | To be estimated |
| $m_2$ | Death rate of $I_c$ class | To be estimated |
| $u_i$<br>($i = 1, 2, 3$) | Control parameter | 0 to 1 |

**Figure 1:**
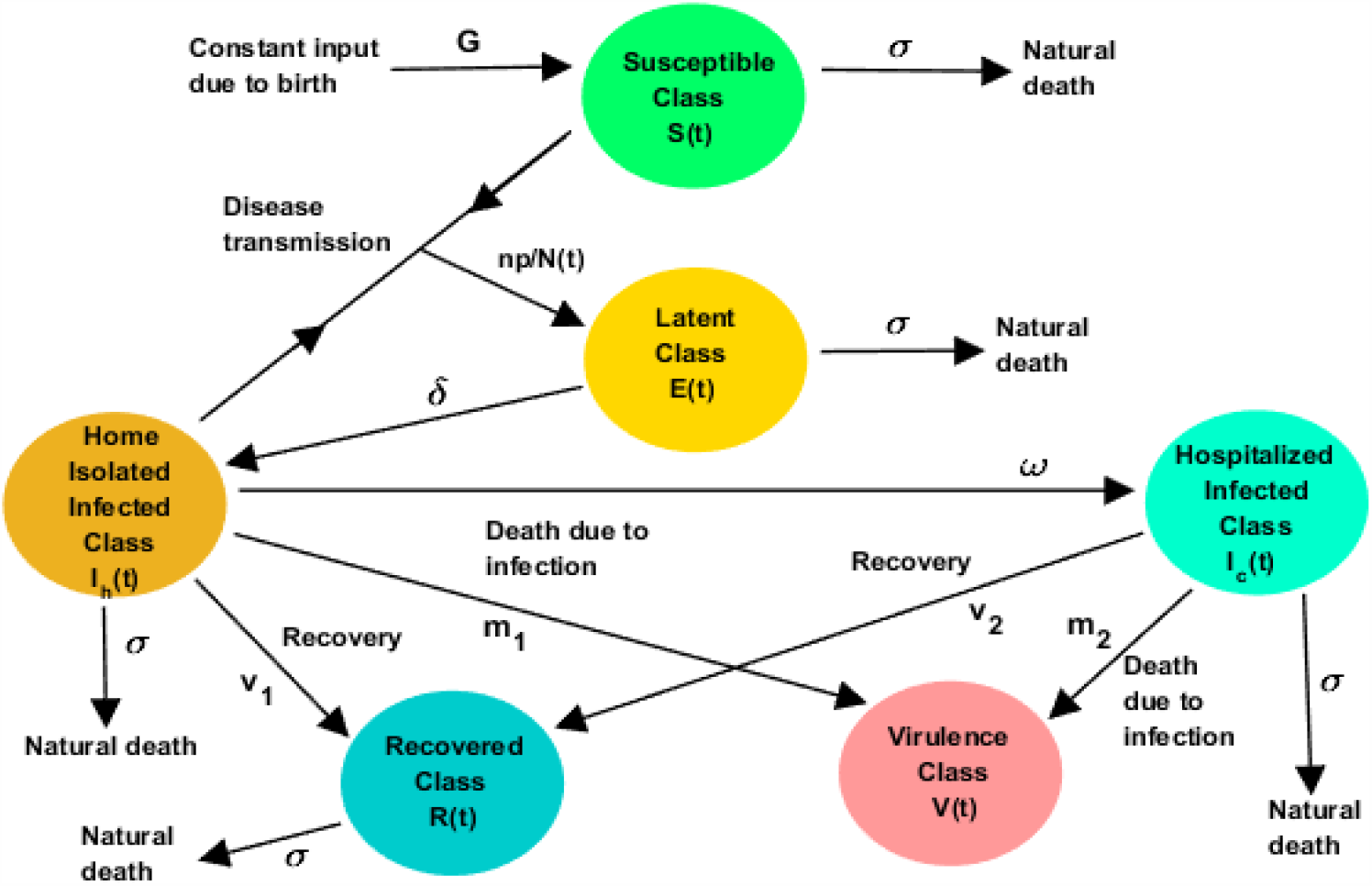
Schematic diagram of the disease progression mechanism considered in the system (2).

We now introduce three constant controls *u*_*i*_ (*i* = 1, 2, 3), 0 ≤ *u*_*i*_ ≤ 1 to the above epidemic model. The control *u*_1_ is introduced to reduce the daily number of contacts due to lockdown (complete or partial). A second control *u*_2_ is introduced to diminish the probability of transmission due to using a face mask and maintaining social distance, individual hygiene, cough etiquette, etc. Thus, (1 − *u*_1_) and (1 − *u*_2_) are the effective daily number of contacts and transmission probability in the presence of the control measures *u*_1_ and *u*_2_. The third and most interesting control *u*_3_ is applied to reduce the death rate and increase the recovery rate by using various repurposing drugs and convalescent plasma therapy. As the effect of the application of repurposing drugs, a fraction of infected people, who earlier succumbed to infection, now recovers and joins *R* class; while the remaining fraction is the member of the disease-related death class, *V*. In fact, without the repurposing drugs, infected individuals will die at a rate *m*_2_ and there will be no extra recovery or a reduction in the death class. Introducing these controls, the system (2) reads

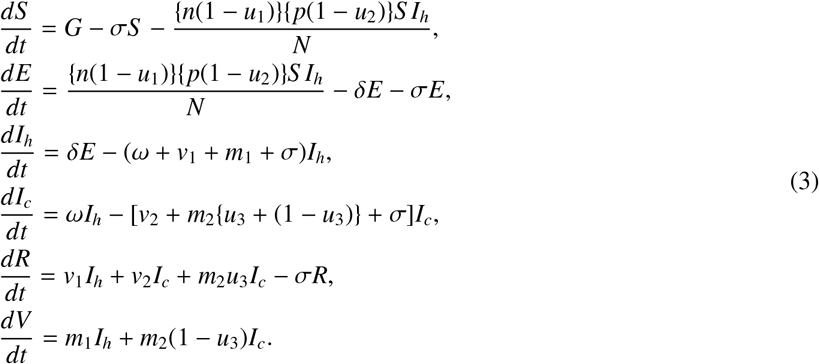

The models (2) and (3) will be analyzed with the initial conditions

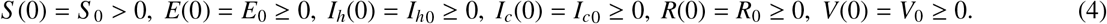

It is to be noted that different sub-cases may be deduced from the model (3) depending on the mitigation measures. For example, an epidemic model of Covid-19 can be deduced for the before lockdown period, when there was no control measure, by setting *u*_1_ = *u*_2_ = *u*_3_ = 0. The model (3) has to be modified with *u*_3_ = 0 if repurposing drugs are not used. A subsystem can be deduced with *u*_1_ = 0 but *u*_2_, *u*_3_ ≠ 0 if there is no lockdown but individual hygiene care and repurposing drugs are present. In the presence of all types of control measures, the variables *u*_1_, *u*_2_, *u*_3_ will have non-zero values. This model, therefore, can capture various lockdown and unlock stages applied in relation to Covid-19 pandemic as well as the recently approved treatment strategy with repurposing drugs.

## 3. Methodologies

We here present the analysis technique of the model and give the stability results of different equilibrium points of the system. First, from biological view point, we show that all solutions of the system (3) are positive and bounded.

### Positivity and boundedness of the solutions

In the sequel, we will use the following well known lemma [46].

Lemma 3.1 *Consider a system* 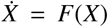, *where* 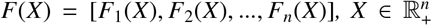, *with initial condition* 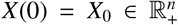. *If for X*_*i*_ = 0, *i* = 1, 2, …, *n*, 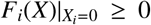, *then any solution of* 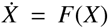 *with given initial condition, say, X*(*t*) = *X*(*t*; *X*_0_) *will remain positive, i*.*e*., 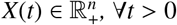.

Proposition 3.1 For initial condition (4), solutions of the system (3) are positively invariant provided *u*_*i*_(*t*) ∈ [0, 1). Moreover, all solutions are uniformly bounded in Γ, where

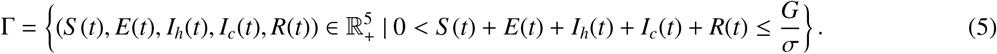

*Proof*. Define *X*(*t*) = (*S* (*t*), *E*(*t*), *I*_*h*_(*t*), *I*_*c*_(*t*), *R*(*t*)). It can be easily seen from (3) that

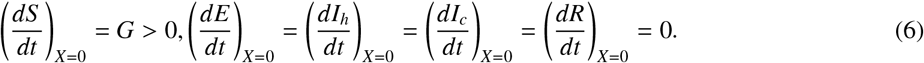

Lemma 3.1 then gives that all solutions of the system (3) starting with the initial condition (4) are positive. Again,

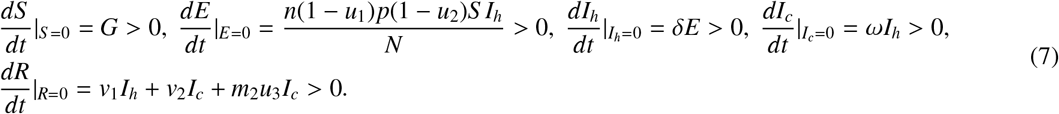

Thus, following [47], 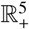 is invariant. Therefore, all solutions of the system (3) with initial condition (4) are positively invariant. To show the boundedness of the solutions of (3), we define

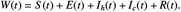

Then we have

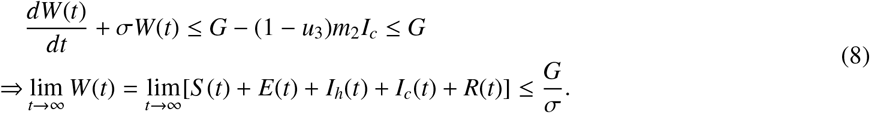

Hence all the solutions of the system (3) are positively invariant and ultimately bounded in the region Γ defined in (5).

□

### Basic reproduction number

The basic reproduction number (*R*_0_) is one of the most important metrics in epidemic theory. It quantifies the condition of disease progression or extinction in a population. More precisely, a disease will grow if *R*_0_ > 1, otherwise it will die out. Here we evaluate the basic reproduction number of the system (3) with the help of next-generation matrix [48]. One can see that the population settles at 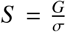 in the absence of infection. The infection subsystem of (3), which describes the production of new infections and changes in the state capable of creating new infections, is given by

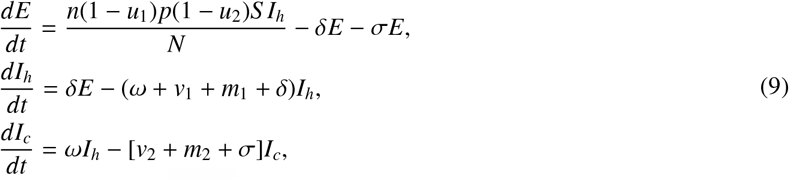

The transmission matrix and transition matrix associated with this infection subsystem (9) in a completely susceptible scenario (when 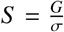) are given by **T** and Σ, respectively, where

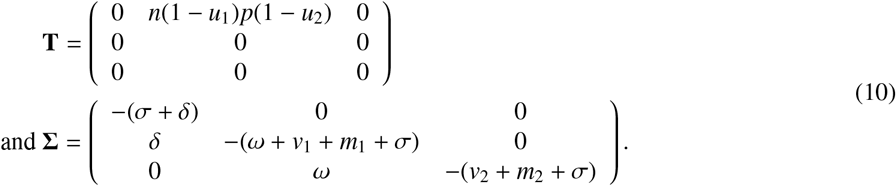

Then the spectral radius of the matrix −**T**Σ^−1^ gives the basic reproduction number of the system (3) and is defined by

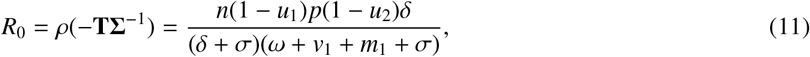

whenever *u*_*i*_∈ [0, 1), *i* = 1, 2. It is straightforward to see that the value of *R*_0_ is higher when there is no control. It is to be noted that whenever *u*_1_ = 1 or *u*_2_ = 1 then the incidence term becomes zero and all subsequent state variables tend to zero as time becomes large, implying that the system becomes infection-free. Therefore, there is no question of the basic reproduction number if *u*_*i*_ = 1, *i* = 1, 2.

### Existence and stability of the equilibria

To determine the asymptotic behavior of the system (3), we find that it has two equilibrium points: (i) the disease-free equilibrium, 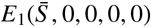 where 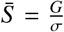, and (ii) the endemic equilibrium 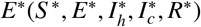. The equilibrium components of *E*^∗^ can be computed as

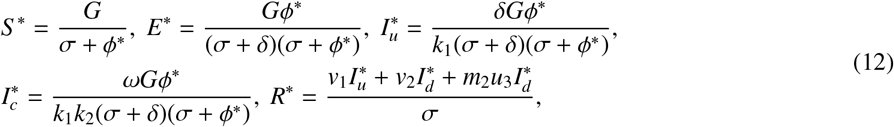

Where

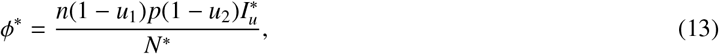

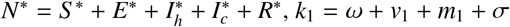 and *k*_2_ = *v*_2_ + *m*_2_ + σ.

Then the Eq. (13) can be reexpressed as

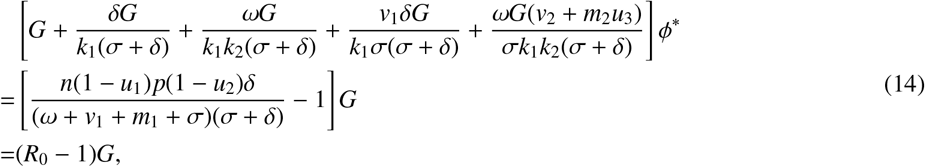

where *R*_0_ is given by (11) with *u*_*i*_∈ [0, 1), *i* = 1, 2. Clearly, there exists a unique interior equilibrium *E*^∗^ of the system (3) whenever *R*_0_ > 1.

We give below the stability results of these equilibrium points.

Theorem 3.1 The disease-free equilibrium *E*_1_ of the system (3) is globally asymptotically stable if *R*_0_ ≤ 1.

*Proof*. Consider the Lyapunov function

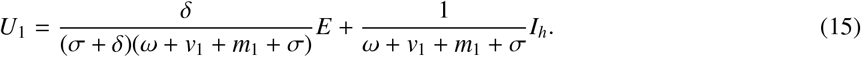

Differentiation of *U*_1_ along the solutions of (3) gives

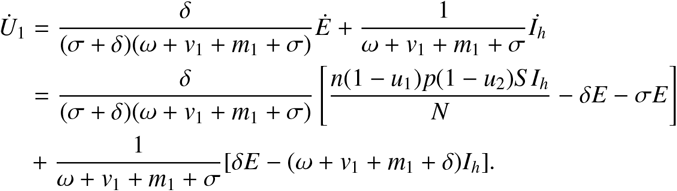

Noting that *S* (*t*) ≤ *S* (*t*) + *E*(*t*) + *I*_*h*_(*t*) + *I*_*c*_(*t*) + *R*(*t*) = *N*(*t*) for all *t* ≥ 0,

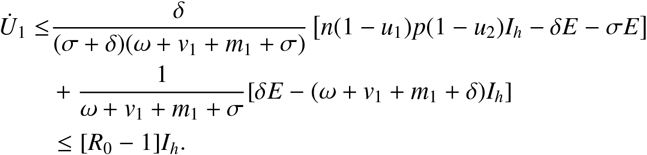

Thus, 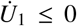 if *R*_0_ ≤ 1. The equality sign occurrs at the disease free equilibrium, *E*_1_. Therefore, using LaSalle’s invariance principle [49], one obtains (*E*(*t*), *I*_*h*_(*t*)) → 0 as *t* → ∞. It gives that lim sup_*t*→∞_ *I*_*h*_(*t*) = 0. Therefore, for any sufficiently small ϵ > 0, there exists a positive constant *M* > 0 such that lim sup_*t*→∞_ *I*_*h*_(*t*) ϵ for all *t* > *M*. From (3), one can have for *t* > *M*,

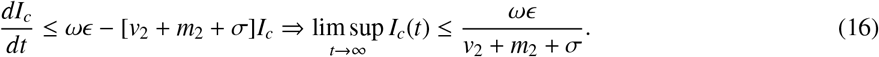

Letting ϵ → 0, we obtain lim sup_*t*→∞_ *I*_*c*_(*t*) ≤ 0. Again, using the fact that lim inf_*t*→∞_ *I*_*h*_(*t*) = 0, lim inf_*t*→∞_ *I*_*c*_(*t*) ≥ 0. Thus, we finally get lim_*t*→∞_ *I*_*c*_(*t*) = 0. In a similar manner, one can show that lim_*t*→∞_ *R*(*t*) = 0 and lim_*t*→∞_ 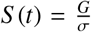. Therefore, all solutions of the system (3) with initial conditions in Γ eventually converge to the disease-free equilibrium *E*_1_ if *R*_0_ ≤ 1. Hence the theorem is proven.

□

Theorem 3.2 The endemic equilibrium *E*^∗^ of the system (3) is locally asymptotically stable whenever it exists, i.e., if *R*_0_ > 1.

*Proof*. The proof is based on the line of [50]. The variational matrix of the system (3) evaluated at *E*_1_ is given by

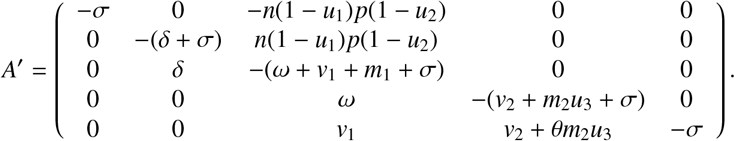

We consider the average per capita daily contacts of an undetected infectious individual, *n*, as the bifurcating parameter and apply the central manifold theorem to determine the local stability of *E*^∗^. The critical value *n* = *n*^∗^ for which *R*_0_ = 1 holds is

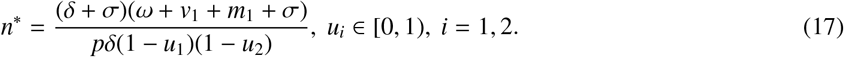

Now let at *n* = *n*^∗^, the Jacobian matrix 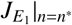 has a right eigenvector *u* = (*x*_1_, *x*_2_, *x*_3_, *x*_4_, *x*_5_)^*T*^ corresponding to the zero eigenvalue, where

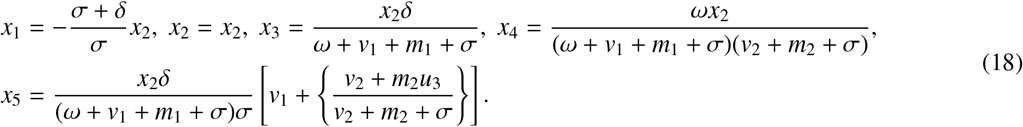

Similarly, a left eigenvector corresponding to the zero eigenvalue of the Jacobian matrix 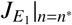 is *w* = (*w*_1_, *w*_2_, *w*_3_, *w*_4_, *w*_5_), where

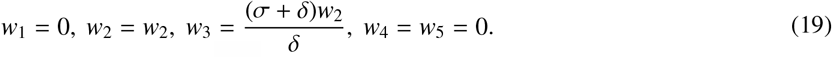

With the transformations *S* = *y*_1_, *E* = *y*_2_, *I*_*h*_ = *y*_3_, *I*_*c*_ = *y*_4_, *R* = *y*_5_, the system (3) can be expressed as

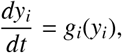

where *g*_*i*_ ∈ C^2^ (ℝ^5^ × ℝ), *i* = 1, ‥, 5. Then the second order partial derivatives of *g*_*i*_ at *E*_1_ are evaluated as

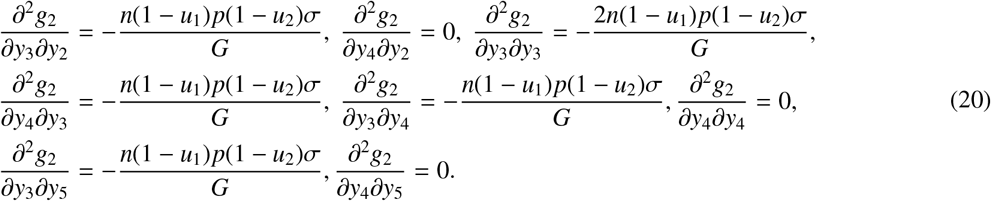

The signs of the quantities α and β evaluated at *n* = *n*^∗^ determine the local stability of the system [50], where

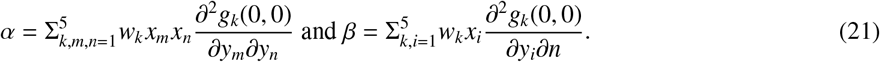

Following the Remark 1 of Theorem 4.1 given in [50], a transcritical bifurcation will occur at *R*_0_ = 1 if α < 0 and β > 0 at *n* = *n*^∗^. Substituting all the values of the second order partial derivative evaluate at *E*_1_ with *n* = *n*^∗^, we then have

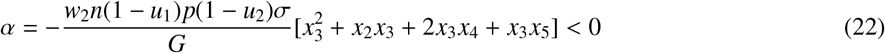

and

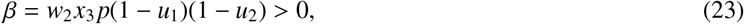

implying a transcritical bifurcation at *R*_0_ = 1. Thus, whenever the endemic equilibrium exists, i.e., when *R*_0_ > 1, it becomes locally asymptotically stable. Hence the theorem is proven. □

## 4. Simulation results: Indian case study

India is the third worst-hit country of the world by the Covid-19 pandemic with a tally of 33,84,576 confirmed cases and 61,695 casualties as of August 27, 2020 (https://covid19india.org). The first confirmed case in the country was reported on January 30 when a student, who returned to home state Kerela from Wuhan, was tested positive. India observed 68 days nationwide lockdown starting from March 25 to May 31, 2020, to restrict the transmission of the coronavirus. Though this lockdown allowed the healthcare providers to take necessary preparations for combating the coronavirus by improving the health care infrastructures, India remained unsuccessful to prevent the spread of infection. Phase wise unlocking process in India started from June 1 [51]. During the first phase of unlocking (June 1-30), religious places, hotels and restaurants were allowed to reopen maintaining the self-distancing protocol. The second phase started from July 1 and to be continued till July 31. During this period more activities in a calibrated manner were allowed. People’s daily life, however, was not normal [52]. Regular domestic and international flights, local transportation, both short-and long-distance train services remain cancelled. All educational institutions, shopping malls, community halls are completely closed since March 25. Night curfew has been continuing since the first day of unlocking. Some states like West Bengal has newly announced complete lockdown twice in a week [53] and Tripura has announced eight consecutive days complete lockdown to check the surge in the number of Covid-19 positive cases during unlocking period [54].

### Data collection

The data for India was collected from the online freely available repository Covid19India.Org (https://www.covid19india.org/). In this website, day-wise data, as well as the cumulative data of corona cases in India and its all states/ union territories are given. We here considered the time series data of confirmed, recovered and deceased cases of India for the study period 1^*st*^ March to 27^*th*^ August 2020, but did not consider the state-wise data.

### Parameter estimations and curve fitting

Study period data was divided into three time windows: 1^*st*^ March to 24^*th*^ March (before lockdown period), 25^*th*^ March to 31^*st*^ May (lockdown period) and 1^*st*^ June to 27^*th*^ August (unlock period). The value of the constant input *G* to the susceptible class through birth is assumed to be equal to the per day new birth 77575 [55]. It is to be mentioned that no immigration or emigration was considered due to international travel restriction. Average life expectancy of an Indian is 66.8 years [55], giving the value 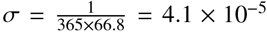. During parameter estimation, it was taken into account that ν_1_ >> ν_2_ and *m*_1_ << *m*_2_. The leaving rate of *E* class is measured by the parameter δ, which is a test-dependent parameter. Covid test in India increased significantly during the later stage compared to the early stage of the epidemic. Accordingly, the value of δ should be lower in the before lockdown stage and higher in the after lockdown period. Further, the effective daily contact *n*(1 − *u*_1_) should be the lowest during the complete lockdown period. On the contrary, the probability (*p*) of disease transmission per contact is expected to be the highest at the initial stage of the epidemic because people were less aware of the importance of individual hygiene in the disease transmission. In the subsequent time, people became more concern about the infection and therefore the effective probability (*p*(1 − *u*_2_)) decreased. The population of India as of January 1, 2020 is considered as the initial value of susceptible population (*S* (0)), which is 1387297452 [55]. The controls were assumed to be zero during the before lockdown period and only two controls (*u*_1_, *u*_2_) were assumed to have non-zero values in the lockdown period as well as unlock period. Therefore, the third control was not used during the study period, but it was used for future prediction. The solution curve of the model system (3) was then fitted with the actual data. In this process, the fminsearch optimization toolbox of Matlab and nonlinear least-square technique were used [56]. We implemented the fminsearch optimization technique in the way described in [57] and have taken the nonlinear least-square method for MATLAB from mathworks repository [58]. The parameter values that best fit the epidemic data with the solution of the controlled system for different time periods are given in Table 2.

**Table 2:** Estimated parameter values for India

| Time period | $n$ | Effective daily contact $n(1 - u_1)$ | $p$ | Effective probability $p(1 - u_2)$ | $\delta$ | $\omega$ | $\nu_1$ | $\nu_2$ | $m_1$ | $m_2$ | $u_1$ | $u_2$ |
| --- | --- | --- | --- | --- | --- | --- | --- | --- | --- | --- | --- | --- |
| 1<sup>st</sup> March- 24<sup>th</sup> March | 2.2199 | 2.2199 | 0.3070 | 0.3070 | 0.4038 | 0.3505 | 0.0134 | 0.0011 | 0.00001 | 0.00571 | 0 | 0 |
| 25<sup>th</sup> March-31<sup>st</sup> May | 3.0875 | 2.1612 | 0.3070 | 0.2763 | 0.4438 | 0.3415 | 0.1554 | 0.0043 | 0.00001 | 0.00145 | 0.3 | 0.1 |
| 1<sup>st</sup> June-27<sup>th</sup> August | 3.7599 | 3.1959 | 0.3003 | 0.1952 | 0.6238 | 0.4205 | 0.1374 | 0.0058 | 0.00001 | 0.0002 | 0.15 | 0.35 |

In Fig. 2, we have presented the curve fitting of the model for the before lockdown period (1^*st*^ March to 24^*th*^ March). It can be observed that fitting is relatively poor for the recovered and deceased classes. The main reason is that there were many inaccuracies in the initial data. In many cases, there were under-reporting of the actual facts, and comorbidities among COVID-19 deaths were also excluded. Such problems related to data, however, resolved later on by the health authority.

**Figure 2:**
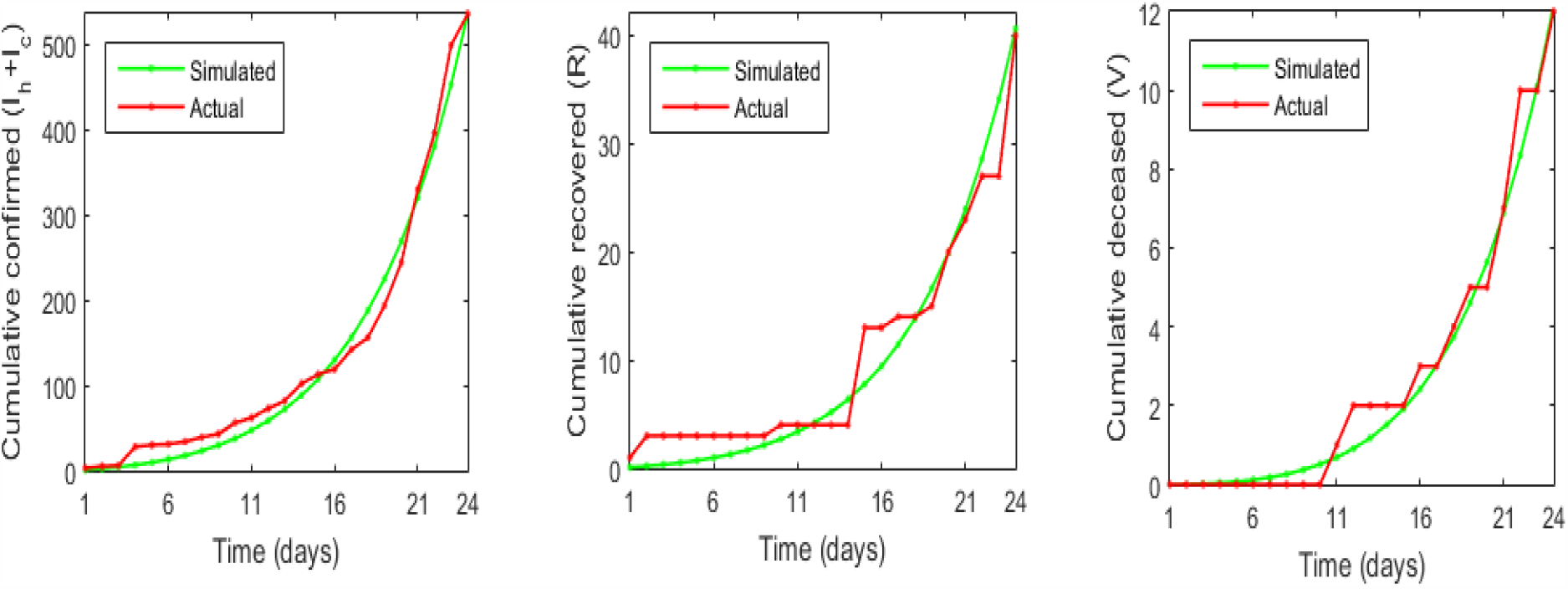
Actual cumulative values (red colour) of confirmed, recovered and death cases in India for the study period March 1 to March 24, 2020 (before lockdown period) are fitted by the solution (green colour) of the system (3) with *u*_1_ = *u*_2_ = *u*_3_ = 0.

In Fig. 3, the actual data between 25^*th*^ March to 31^*st*^ May (lockdown period) and the data of the simulated results of the controlled system (3) with fixed values of *u*_1_ = 0.3, *u*_2_ = 0.1 and *u*_3_ = 0 are shown. A similar figure for the time period 1^*st*^ June to 27^*th*^ August (unlock period) with fixed control *u*_1_ = 0.15, *u*_2_ = 0.35 and *u*_3_ = 0 are presented. These curves show good fitting of actual and simulated data.

**Figure 3:**
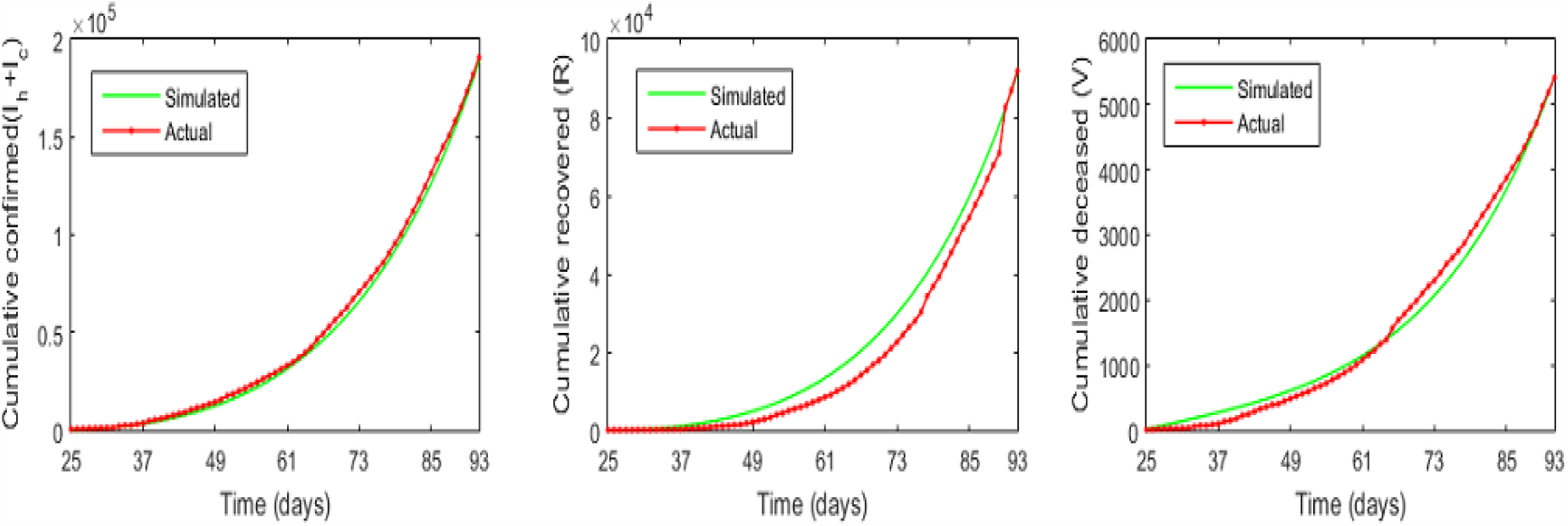
Actual cumulative data (red colour) of confirmed, recovered and death cases in India for the study period March 25 to May 31, 2020 (lockdown period) are fitted by the solution (green colour) of the system (3) with fixed controls *u*_1_ = 0.3, *u*_2_ = 0.1 and *u*_3_ = 0.

**Figure 4:**
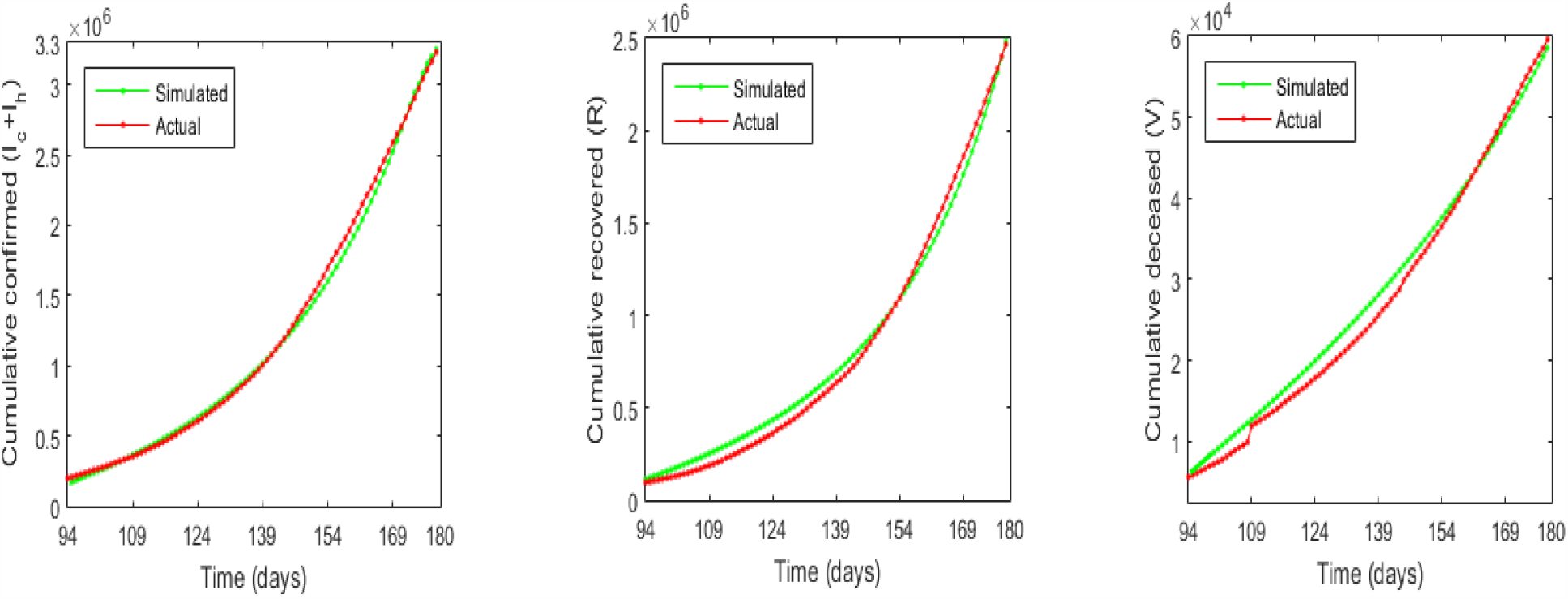
Actual cumulative values (red colour) of confirmed, recovered and death cases in India for the study period 1^*st*^ June to 27^*th*^ August, 2020 (unlock period) are fitted by the solution (green colour) of the system (3) with fixed controls *u*_1_ = 0.15, *u*_2_ = 0.35 and *u*_3_ = 0.

### Effect of repurposing drugs

Here we explore the effect of repurposing drugs in treating covid-19 patients. Clinical results show that at least one-third of critically ill covid patients can survive by administrating the repurposing drug dexamethasone [23, 24]. Different countries have given approval of such life-saving drugs. The Government of India gave approval of using flavipiravir on July 23 [59] for the treatment of Covid-19 patients. Some states of India are supposed to get the first batch of coronavirus drug remdesivir for the use of covid patients very soon [60]. In addition, the life of some covid patients can be saved by applying convalescent plasma therapy also [30]. We here used three different values of the control parameter *u*_3_ with the parameter values of the unlock period and demonstrated how repurposing drugs can reduce the epidemic burden. In Fig. 5, the green colour curve represents the cumulative values of the confirmed, recovered and death cases in the next one month if the repurposing drugs are not used. The other three curves in these figures represent similar cumulative numbers if these drugs are used. The right figure shows that the number of deaths after one month will be 97243 if no repurposing drug is used (i.e., *u*_3_ = 0). This number would be 92449 if the repurposing drugs are used with controlling efficacy *u*_3_ = 0.3. Thus, 4794 deaths can be avoided in the next one month (August 28 to September 26) when *u*_3_ takes value 0.3. This number will be 2353 and 7303 corresponding to the drug efficacies *u*_3_ = 0.2 and *u*_3_ = 0.4, respectively.

**Figure 5:**
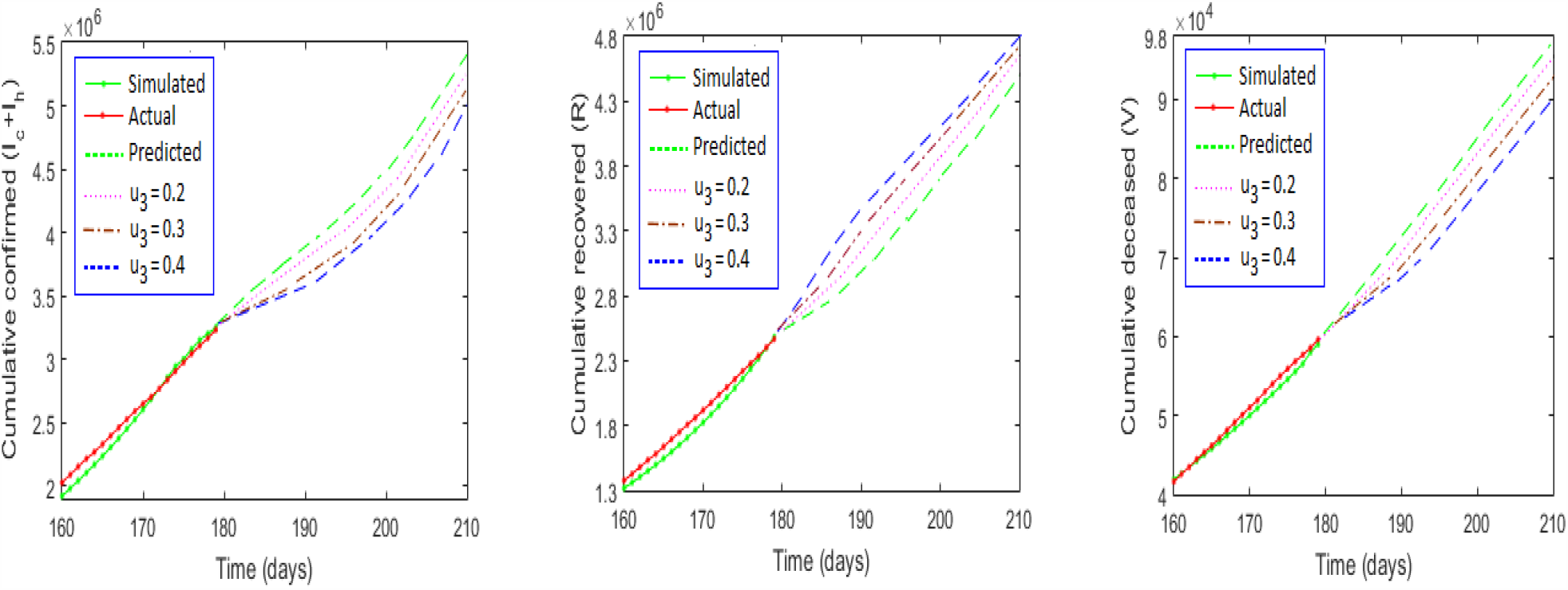
Predicted cumulative Covid-19 confirmed, recovered and death cases in India for the next one month (August 28 to September 26) for different values of the control *u*_3_. Here 180 days implies August 27, 2020, and 210 days implies September 26, 2020. The predicted results with *u*_3_ = 0 is represented by the green dashed line in each figure. The red, brown and blue colours represent the same for *u*_3_ = 0.2, *u*_3_ = 0.3 and *u*_3_ = 0.4, respectively. The cumulative number of deaths as of September 26 when *u*_3_ = 0 are 97,243 and the same for *u*_3_ = 0.3 are 92.449, implying 4,794 fewer deaths in one month. Parameters are as in Table 2 (unlock period).

## 5. Discussion

A highly contagious respiratory disease Covid-19, which originated from Wuhan city, China at the end of 2019, spread over the entire globe very rapidly. No country was able to protect its people from the invasion of this virus. The world has never seen such devastating effect of any pathogen in the last 100 years. Though WHO, for the sake of better management and accelerated preparedness, announced 2019-nCoV outbreak as of public health emergency of international concern (PHEIC) and later on declared it as a pandemic, blocking the spread of this novel virus SARS-CoV-2 remained unsuccessful.

Mathematical models are vital in understanding the disease dynamics and may be used as an important tool in projecting the epidemic burden in the near future. Such estimation may be used to design effective control strategies and necessary facility development to combat the disease. Lots of mathematical models have been proposed in the meantime to forecast about the timing of epidemic peak, number of cases to be expected per day and the expected duration of the epidemic. It is, however, to be mentioned that prediction is not enough, we should at the same time prescribe how to optimize the use of limited resources for better management of the epidemic. In this process, one can compare the outcomes of the available treatment and the infection controlling strategies to prescribe the best possible option so that the health administrators can take the right decision to save the life of infected subjects and limit the epidemic. In this work, we have proposed a mathematical model that divides the population of a covid-infected country into five disjoint classes depending on the health status of its subjects and then compared the outcomes of different controlling strategies as well as various treatment options currently available. Use of repurposing drugs in the mathematical model to study the Covid-19 epidemic burden is unique and has not been considered in earlier studies.

Basic reproduction of an epidemic model usually delineates the stability and existence of a disease-free state from its endemic state. More precisely, if the basic reproduction number *R*_0_ is greater than unity then an epidemic can spread, otherwise epidemic dies out. We determined the basic reproduction number of our system by next-generation matrix and it showed that the classical properties of the basic reproduction number remained intact for Covid-19 epidemic. In the case of India, the basic reproduction number significantly reduced during the lockdown period, however, it remained almost the same during the unlock period (Fig. 6(a)). To contain the epidemic, the value of *R*_0_ has to be brought down below 1 through NPIs in absence of vaccine. It is worth mentioning that the value of the nonpharmaceutical control parameters must be significantly high in this case. Fig. 6(b) shows that any combination of the control parameters *u*_1_ and *u*_2_ that lies above the curve *R*_0_ = 1 for the unlock parameters can eliminate SARS-CoV-2 virus from the system. India is trying hard to prevent the transmission of coronavirus by implementing NPIs. People have become more aware of this deadly virus and following the statutory instructions more sincerely than before. Some state governments even have imposed punitive measures for flouting covid controlling norms [61, 62, 63]. Community involvement may certainly help to push the point (*u*_1_, *u*_2_) = (0.15, 0.35) in the region *R*_0_ > 1 to the region *R*_0_ < 1, resulting in the elimination of infection.

**Figure 6:**
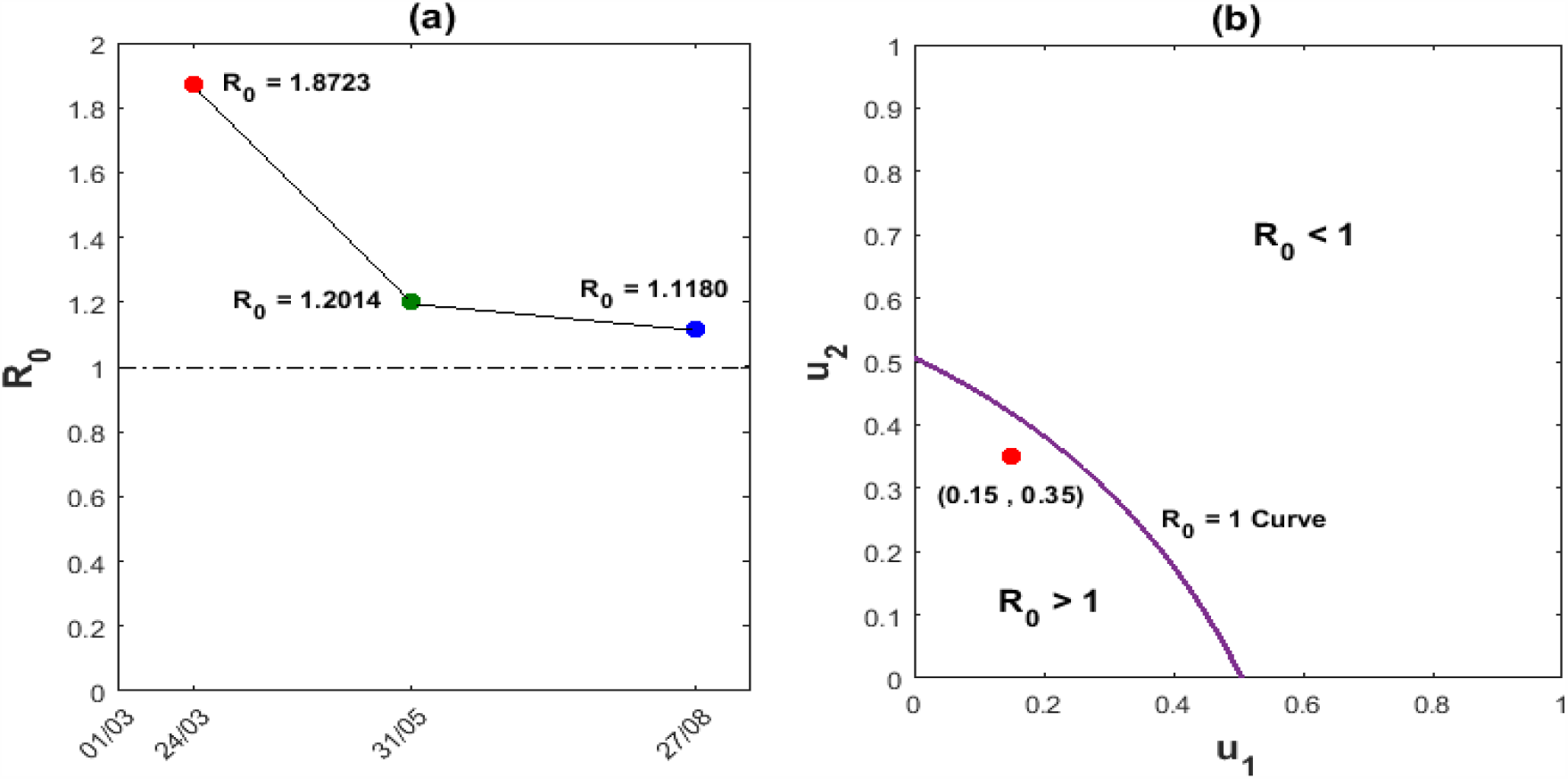
(a) Variation in the basic reproduction number, *R*_0_, of the Covid-19 epidemic in India for the study period March 1 to August 27, 2020. It shows that *R*_0_ decreases from its value 1.8723 as of March 24 to 1.1180 as of August 27. March 24 is the lockdown starting date and May 31 is the lockdown ending date for India. The daily positive cases will decline from its current value once *R*_0_ goes below the dashed line *R*_0_ = 1. (b) The curve *R*_0_ = 1 separates the infection-free state (*R*_0_ < 1) from the endemic state (*R*_0_ > 1) in the plane of control parameters *u*_1_, *u*_2_. The present estimated values of *u*_1_ and *u*_2_ are marked with a red dot. Parameters are as in the Table 2 with unlock case.

**Figure 7:**
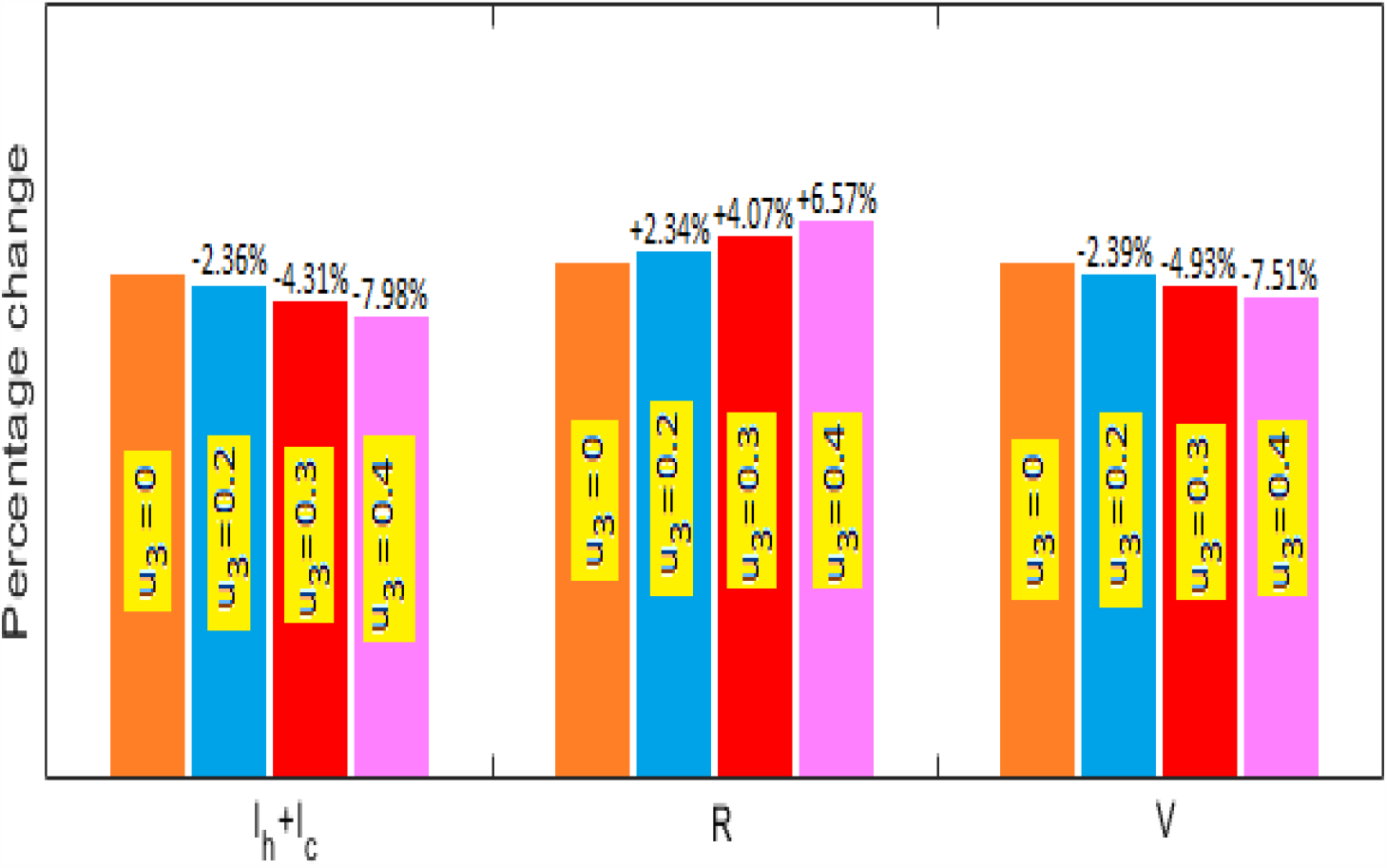
Percentage change in the projected numbers of positive, recovered and death cases as of September 26 due to the use of repurposing drug therapy with different control efficacies. The change is calculated with their corresponding values of August 27. It shows that the use of repurposing drugs will reduce the projected deaths by 2.39%, 4.93% and 7.51% when *u*_3_ takes values 0.2, 0.3 and 0.4, respectively. Similar changes may be observed in the cumulative infected cases (*I*_*h*_ + *I*_*c*_). An increase in the recovery class (*R*) is also observed and the percentage change is indicated by a positive sign.

Some repurposing drugs have brought new hopes for critically ill Covid-19 patients. It can significantly reduce the covid related deaths and increase the recovery number. Our simulation results show that recovery can be increased by 6.57%, and death & positive cases can be reduced, respectively, by 7.51% & 7.98% (corresponding to the control parameter *u*_3_ = 0.4) within a month by using repurposing drugs. This study shows that containment of coronavirus by NPIs only is a hard task for India but definitely not impossible. The use of repurposing drugs is very successful in saving lives and reducing the number of infectives. The combined use of NPIs and repurposing drugs may significantly improve the overall Covid-19 epidemic burden. It is to be mentioned that we have used fixed control in our study, one can, however, consider these controls as time-varying in finding optimal control strategies to reach out to the target. In this case, the objective should be to minimize the number of individuals to all infection-related classes, the decease related deaths and the related cost.

To get rid of this Covid-19 pandemic, we have to be united and fight together to eliminate the infection from each corner of the world. We have to keep in mind that the disease started spreading from a market of Wuhan and eventually spread throughout the globe. In combating the global epidemics, collaboration among industry, academia, and public health entities are essential [64].

## Data Availability

The data used here is freely available at https://www.covid19india.org/

## Acknowledgment

Research of C.M. is supported by CSIR, India [F. 09/096(0865/2016-EMR-I)] and Research of N.B. is supported by SERB, Ref. No.: MSC/2020/000020.

